# Mapping the intersection of social status and comorbidity in knee osteoarthritis: a WOMAC-based study

**DOI:** 10.1101/2025.05.03.25326939

**Authors:** Mohoshina Karim, Md Delwar Hossen, Tasrima Trisha Ratna, Fatema Priyanka, Ilat-E-Mees Subah, Umme Salma, Rahnuma Hossain Twasin, Joynal Abedin Imran, Shahriar Hasan, Marzana Afrooj Ria

## Abstract

Knee osteoarthritis (OA) is a disabling joint condition that leads to extreme mobility and quality of life impairment, particularly among older adults. This study aimed to investigate the socio-demographic factors and comorbid conditions influencing the severity of symptoms of knee OA using the Western Ontario and McMaster Universities Osteoarthritis Index (WOMAC). Data were derived from 622 patients across 9 months from the major healthcare facilities of Dhaka. Age, sex, educational status, obesity, diabetes mellitus, and cardiovascular disease (CVD) were predictors for the severity of symptoms of OA of the knee, the study claimed. Female participants were more prone to have severe symptoms compared to males, and those who were more than 70-years-old were at greater risk of severe symptoms. Low educational status, obesity, diabetes mellitus, and CVD were also predictors for severe OA of the knee. Age (p<0.001), obesity (p<0.001), and diabetes (p<0.001) were the best predictors of severity of symptoms based on the multinomial logistic regression analysis. The findings from the study highlight the complex etiology of OA of the knee and the need for integral healthcare measures that address both the socio-economic and the physical determinants. Focused interventions need to be employed, particularly for high-risk groups such as the elderly, women, and the comorbid, to minimize the incidence of OA of the knee and maximize the outcomes for patients in settings such as that of Bangladesh, where resources may not be available.

## Introduction

Osteoarthritis (OA) is a degenerative joint disease that gives rise to stiffness, pain, and loss of mobility, which predominantly occurs among older adults [1]. OA is one of the main contributors to worldwide disability, with an upward increase in incidence due to aging populations, as well as rising obesity rates [2]. The condition poses a huge healthcare burden since it negatively affects the quality of life of an individual, particularly of the knee joints [3].

Articular cartilage degeneration, subchondral bone remodeling, synovial inflammation, and osteophyte formation characterize Knee OA [4,5]. Pain, stiffness, and functional disability result from these pathological processes and have a significant effect on physical function and overall quality of life [6,7]. The progression of OA encompasses a multifaceted interaction between mechanical stress, inflammation, and oxidative damage [8], and worsening symptoms and limitations of function [9].

Knee OA among older people leads to pain, reduced mobility, limitation of functions, loss of independence, and also to impaired social participation [10,11]. Estimated prevalence of knee OA among adults is 7.3% for Bangladesh [12,13], which is a public health concern. Osteoarthritis (OA) imposes high socio-economic burdens on both people and health care systems via increased consumption of healthcare, disability, and loss of productivity [9,14]. The pocket cost of OA not only burdens personal economies of the patients but also public healthcare services [15,16].

Age is also a significant influence on severity and progression of OA knees, where conditions are more severe and degenerate more quickly among older adults [17]. There is also greater vulnerability to knee OA among women compared to men, where there is greater incidence and more severe conditions, particularly post menopause [18,19]. Hormonal deficiency, for example, estrogen deficiency, is significant in aggravating vulnerability among women, for instance, for OA knees [20]. Also, those who are not literate have greater severity of OA since they are not aware of the condition, or they do not reach healthcare services due to limited literacy [21].

Obesity hugely magnifies knee OA by increasing joint mechanical stress and triggering inflammatory processes systemically, speeding disease progression and worsening symptoms [22]. Diabetes mellitus also worsens knee OA by increasing joint pain, impaired function, and inflammation [23]. Diabetic patients also experience more severe symptoms, reduced mobility, and worse outcomes compared to non-diabetic patients [23].

Western Ontario and McMaster Universities Osteoarthritis Index (WOMAC) is a widely used and validated measure of pain, stiffness, and physical function among people with OA of the knee [24,25]. Aligning to its three subscales—pain, stiffness, and physical function—it reports both severity of symptoms and functional disability [24,25]. WOMAC is an important measure for monitoring follow-up improvement in OA of the knee, and also for assessing outcome of treatment [24,25].

Given the significant impact created by osteoarthritis (OA) of the knees among elderly people, this study aimed to examine the influence of socio-demographic factors and comorbid conditions on OA severity in Bangladesh. The increasing prevalence of OA of the knees among elderly people, coupled with high obesity, diabetes, and cardiovascular disease prevalence, underscores the need to establish evidence for factors that affect disease severity. By application of the WOMAC index, this study aimed to provide significant contributions regarding how sociodemographic factors such as gender, educational attainment, income, and age, and also factors related to an individual’s health such as obesity and diabetes, affect severity in OA of knees. The contributions of the study will towards strengthening target interventions and also healthcare delivery within low-resource settings such as Bangladesh.

## Materials and methods

### Study population

This research utilized cross-sectional descriptive design, in three prominent healthcare centers in Dhaka, Bangladesh: Dhaka Medical College Hospital (DMCH), National Institute of Traumatology and Orthopaedic Rehabilitation (NITOR), and Bangladesh Institute of Research and Rehabilitation in Diabetes, Endocrine and Metabolic Disorders (BIRDEM). The hospitals have heterogeneous patient populations with specialized services, thus making them suitable for study. Data were collected over 9 months from April 2024 to December 2024. The trial included individuals with knee OA, regardless of gender, older than 60 years at study initiation. All study participants met diagnostic guidelines for OA in knees by the American College of Rheumatology (ACR), including both clinical and radiographic criteria for OA. Participants who were severely ill, mentally impaired, or lack of ability to provide informed consent were excluded. Individuals with other inflammatory joint diseases (e.g., rheumatoid arthritis, ankylosing spondylitis, psoriasis, gout) and with metabolic or congenital joint diseases were not included to prevent confounding. Participants with histories of orthopedic surgeries were excluded in order to prevent potential biases resulting from such intervention.

### Sampling process

A simple random sampling approach was employed to recruit participants. A total of 622 participants who met the inclusion criteria were selected. Participants who provided written consent were recruited. When a participant refused, the next participant in the line who could be recruited was approached. This avoided sampling bias and minimized non-response bias, hence increasing the validity of the research.

### Survey instruments

Data were gathered with a Bengali validated English version of self-administered questionnaires by means of face-to-face interviews with the participants. Reliability of the instrument was confirmed by a Cronbach’s alpha of 0.95, indicating strong internal consistency. Before filling in questionnaires, data collectors informed respondents about the study purpose and that participation is voluntary. The questionnaire included components on socio-demographic and comorbidity information’s, and the Western Ontario and McMaster Universities Osteoarthritis Index (WOMAC). WOMAC is widely applied, validated, and used to measure pain, stiffness, and physical function in individuals with knee osteoarthritis [24,25].

### Data analysis

After data collection, an MS excel sheet was developed. All data were handled in the MS excel sheet; thereafter, Stata version 16 was utilized for analytical exploration. Chi-square analysis was used to calculate descriptive statistics of socio-demographic features, comorbidities, and frequency of WOMAC outcome. Further, multinomial logistic regression analysis was used in analytical part to test association between socio-demographic and comorbidities with WOMAC scale. The WOMAC scale was scored on the basis of None (0), Mild (1), Moderate (2), Severe (3), and Extreme (4) on a 0-4 scale. Having a score out of a possible 0-20 for pain, 0-8 for stiffness, and 0-68 for physical function, all the values for the subscales are summed. The total score 96. To streamline the analysis, we have recategorized the grading. The recategorized grading is given below: Moderate (0 - 48), Severe (49 - 72) and Extreme (73+). Both unadjusted and adjusted regression models were used to analyze the impact of individual explanatory variables.

### Socio-demographic and health related characteristics

The explanatory variables in this study are categorized as follows: Age group (60 to 69 years vs 70 years and above), Gender (Male vs Female), Marital Status (Married vs Unmarried), Education (Illiterate, Primary, Secondary and above), Monthly family income (25,000 and below, 25,001 to 49,999, 50,000 and above), Financial independence (Independent vs Dependent), Obesity (Yes vs No), Diabetes (Present vs Absent), and cardiovascular disease (Present vs Absent). The primary outcome variable in this study was the WOMAC score, which was used to assess pain, stiffness, and functional limitations in knee OA patients.

### Ethical consideration

Ethical approval for this study was obtained from the Institutional Review Board (IRB) of the National Institute of Traumatology and Orthopaedic Rehabilitation (NITOR/PT/93/lRB/2024/02). Informed consent was obtained from all participants prior to their inclusion in the study.

## Result

Figure 1 illustrates the distribution of WOMAC scores in the 622 participants with 9.1% reported moderate osteoarthritic pain in their knees, 53.8% reported severe pain, and 36.9% faced extreme symptom in their knees.

**Fig 1.**
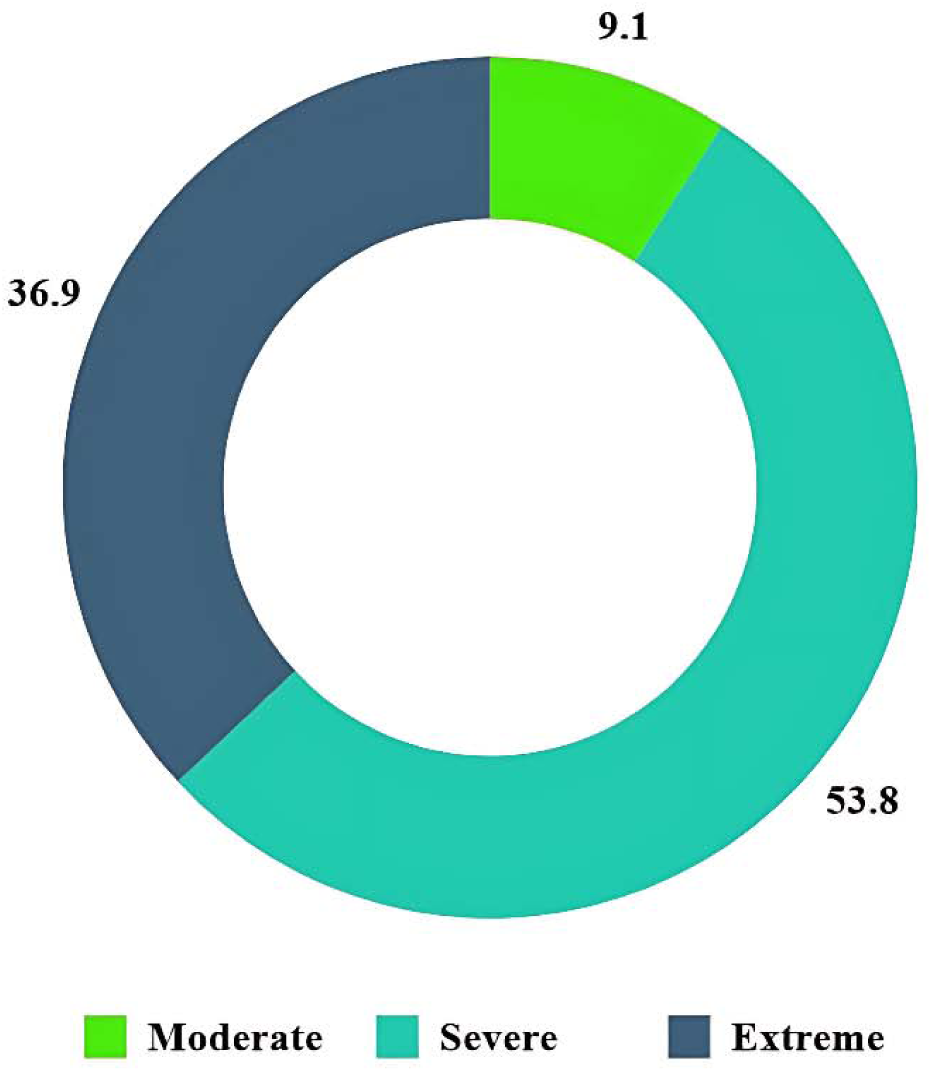
Distribution of WOMAC score among the participants (*n* = 622)

The study also found that there were meaningful relationships between socio-demographic variables and WOMAC scores (Table 1). Age was one such factor, whereby respondents older than 70 years reported higher percentages of severe (55.1%) and extreme (37.3%) symptom compared with respondents between 60 and 69 years. This implies that older individuals have more severe symptoms of knee osteoarthritis. Gender was also found to have an impact since more extreme symptoms were experienced by women (40.6%) compared to men (31.1%), suggesting that women tend to have more severe symptoms of knee OA.

**Table 1.** Association of sociodemographic status with WOMAC score (*n* = 622)

| Variables | Category | Severity |  |  | Chi-square | p-value |
| --- | --- | --- | --- | --- | --- | --- |
|  |  | Moderate (%) | Severe (%) | Extreme (%) |  |  |
| Age | 60 to 69 years | 19 (15.4) | 60 (48.8) | 44 (35.8) | 7.39 | 0.025<sup>b</sup> |
|  | 70 and above years | 38 (7.6) | 275 (55.1) | 186 (37.3) |  |  |
| Gender | Male | 32 (13.6) | 130 (55.3) | 73 (31.1) | 11.89 | 0.003<sup>b</sup> |
|  | Female | 25 (6.5) | 205 (53) | 157 (40.6) |  |  |
| Marital Status | Married | 45 (9.9) | 251 (55) | 160 (35.1) | 3.02 | 0.221 |
|  | Unmarried | 12 (7.2) | 84 (50.6) | 70 (42.2) |  |  |
| Education | Illiterate | 5 (2.6) | 90 (47.4) | 95 (50.0) | 36.38 | 0.000<sup>c</sup> |
|  | Primary | 18 (8.6) | 114 (54.5) | 77 (36.8) |  |  |
|  | Secondary and above | 34 (15.2) | 131 (58.7) | 58 (26.0) |  |  |
| Monthly family income<sup>a</sup> | 25,000 and below | 17 (8.1) | 108 (51.4) | 85 (40.5) | 9.11 | 0.058 |
|  | 25,001 to 49,999 | 22 (7.9) | 148 (53) | 109 (39.1) |  |  |
|  | 50,000 and above | 18 (13.5) | 79 (59.4) | 36 (27.1) |  |  |
| Financial independence | Independent | 32 (12) | 140 (52.4) | 95 (35.6) | 4.48 | 0.106 |
|  | Dependent | 25 (7) | 195 (54.9) | 135 (38.0) |  |  |
| <sup>a</sup>Monthly family income was collected in BDT (1 USD = 121.38 BDT); <sup>b</sup>Significance level (<0.05); <sup>c</sup>Significance level (<0.001) |  |  |  |  |  |  |

Education level was associated with symptom severity, with the highest percentage of extreme symptoms reported by participants with illiteracy (50%), followed by respondents with primary education (36.8%) and secondary education or higher (26%). This implies that less education is related to more severe symptoms. Marital status bore no association with WOMAC scores. Likewise, monthly income from families showed little relation to WOMAC scores, although poorer respondents (25,000 or less) reported an even higher percentage of extreme symptoms (40.5%). Financial independence was similarly not related to WOMAC scores. However, dependent individuals reported a higher percentage with extreme symptoms (38%) compared to independent individuals (35.6%), indicating that financial independence may have some but limited impact on symptom severity.

Table 2 reflects considerable correlations between diabetes, cardiovascular disease, and obesity with WOMAC scores. Participants with obesity reported higher percentages of extreme (42.3%) and severe (52.3%) symptoms compared to non-obese individuals, who indicated a lower percentage of extreme (26.5%) symptoms. This reflects that obesity considerably augments the severity of symptoms of knee osteoarthritis. In the same manner, diabetes was found to have considerable correlations with WOMAC scores. Participants with diabetes reported higher percentages of extreme (37.2%) and severe (54.7%) symptoms compared to non-diabetic individuals, who indicated a considerably lower percentage of extreme (32.3%) symptoms. This reveals that diabetes considerably enhances the severity of symptoms of knee osteoarthritis. Cardiovascular disease (CVD) was also found to have considerable correlations with WOMAC scores. Participants with CVD reported a higher percentage of extreme (44.7%) symptoms compared to non-CVD, who indicated a smaller percentage (23.7%) of extreme symptoms. This reflects that cardiovascular disease further adds to the severity of symptoms of knee osteoarthritis.

**Table 2.** Association of comorbidities with WOMAC Score (*n* = 622)

| Variables | Category | Severity |  |  | Chi-square | p-value |
| --- | --- | --- | --- | --- | --- | --- |
|  |  | Moderate (%) | Severe (%) | Extreme (%) |  |  |
| Obesity | Yes | 22 (5.4) | 215 (52.3) | 174 (42.3) | 29.14 | 0.000<sup>a</sup> |
|  | No | 35 (16.6) | 120 (56.9) | 56 (26.5) |  |  |
| Diabetes | Present | 48 (8.1) | 323 (54.7) | 220 (37.2) | 15.64 | 0.000<sup>a</sup> |
|  | Absent | 9 (29) | 12 (38.7) | 10 (32.3) |  |  |
| Cardiovascular disease | Present | 28 (7.1) | 190 (48.2) | 176 (44.7) | 28.50 | 0.000<sup>a</sup> |
|  | Absent | 29 (12.7) | 145 (63.6) | 54 (23.7) |  |  |
| <sup>a</sup>Significance level (<0.001) |  |  |  |  |  |  |

Table 3 shows the odds ratios (OR) for sociodemographic variables and for comorbidities in relation to severe and extreme knee osteoarthritic pain. Participants aged 70 years and older had more than four times higher odds for experiencing both severe (OR = 4.79, 95% CI: 2.18-10.50, p = 0.000) and extreme (OR = 4.23, 95% CI: 1.84-9.71, p = 0.001) symptoms compared to respondents 60-69 years old. These outcomes were similar in both unadjusted models as well as in adjusted ones, thereby emphasizing that age was a determining factor for symptom severity.

**Table 3:**
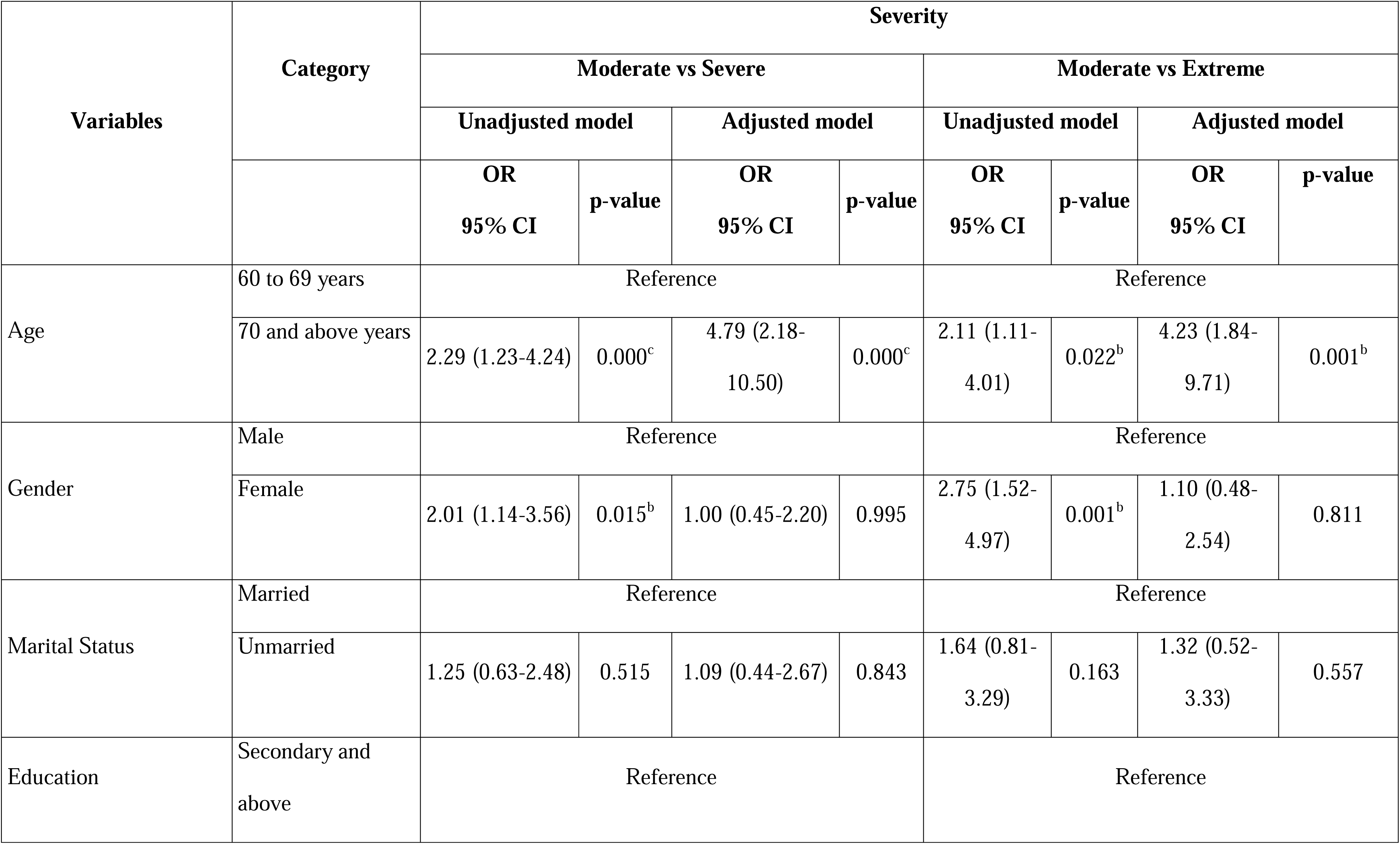
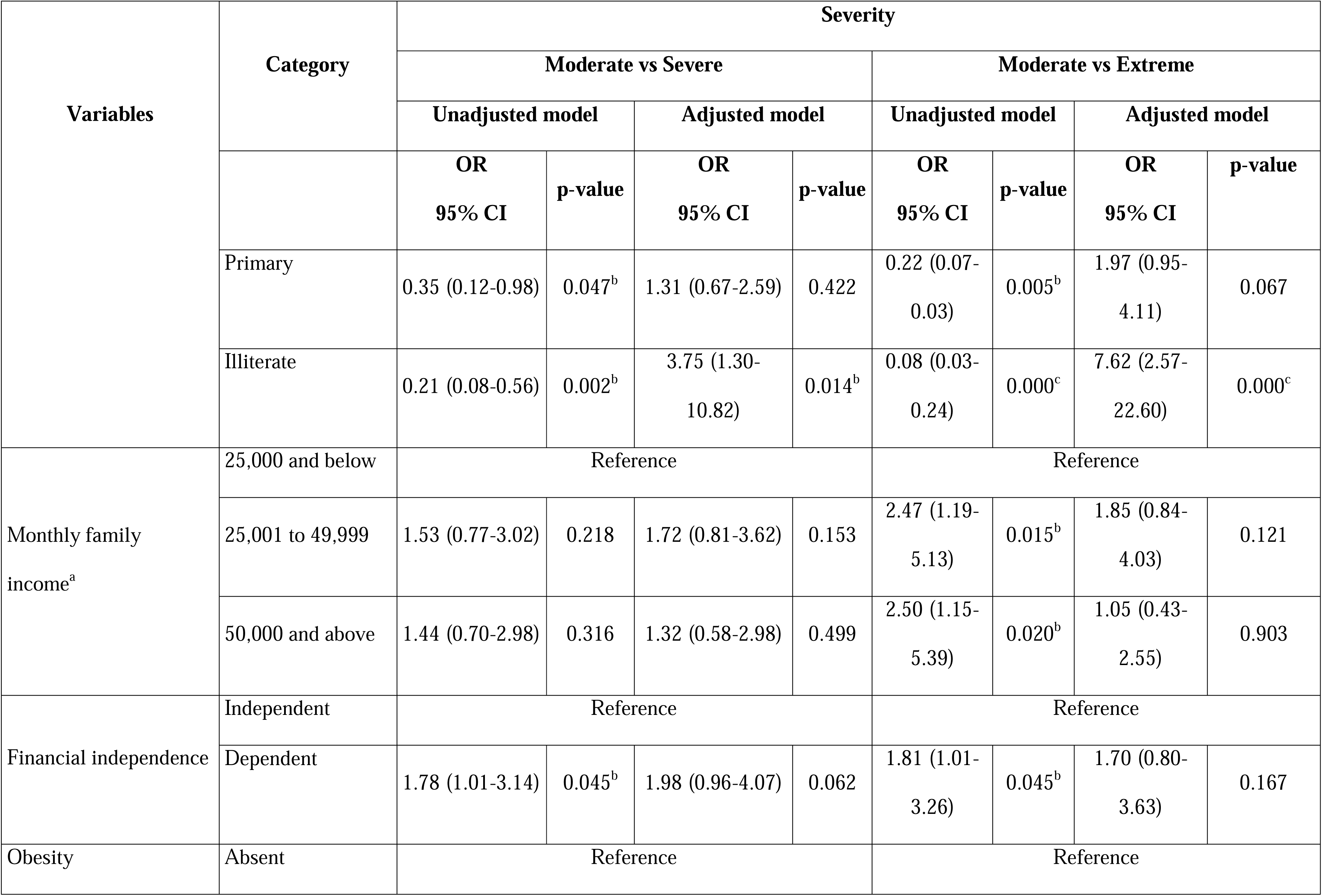
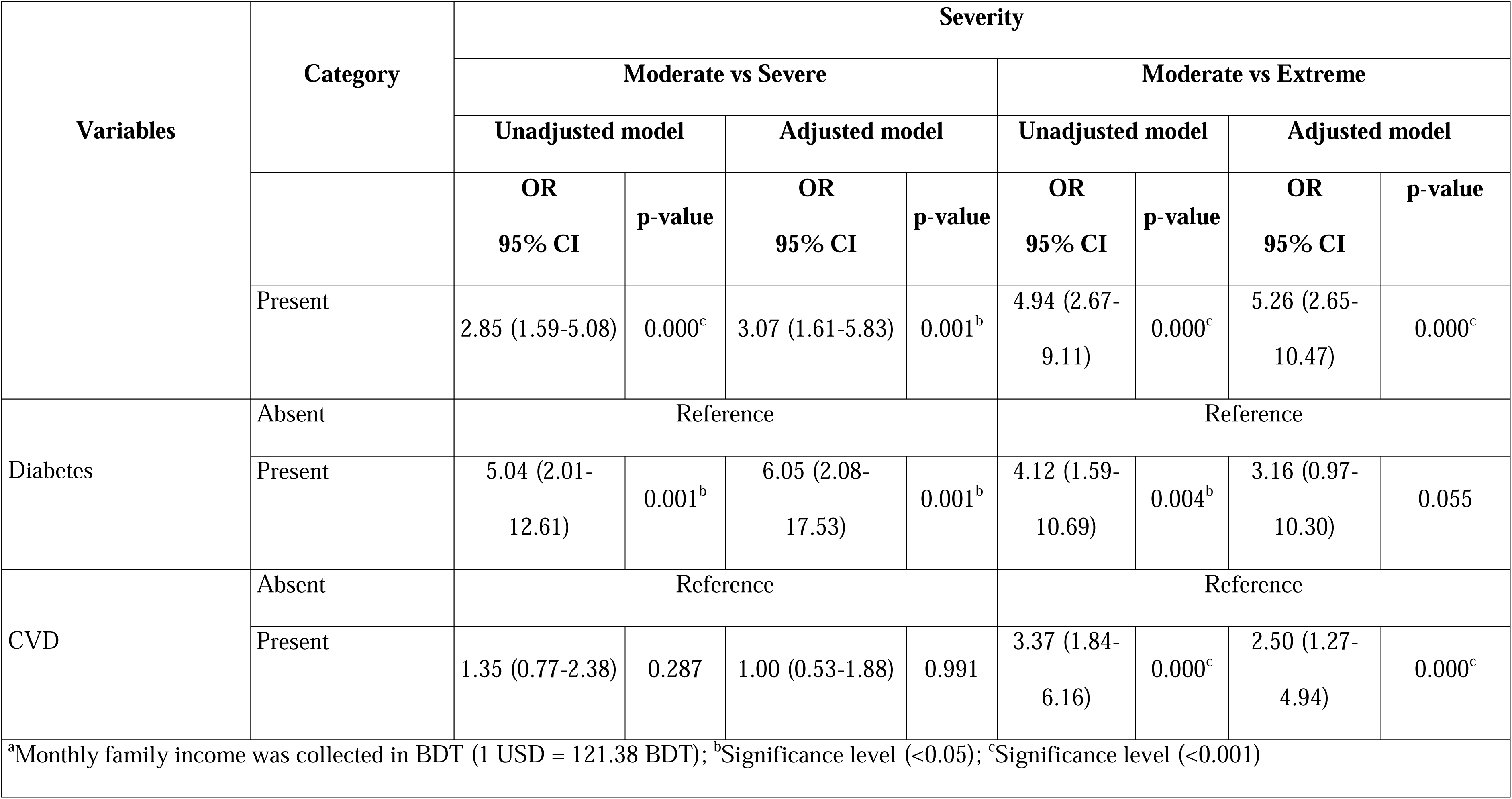
Logistic regression analysis of sociodemographic status and comorbidities with WOMAC Score (Multinomial Logistic regression)

Female respondents had 2.75 times higher odds for experiencing extreme symptoms (OR = 2.75, 95% CI: 1.52-4.97, p = 0.001) compared to males, thus pointing towards gender as an influencing factor for severity in OA symptoms. Participants who were illiterate had 7.62 times higher odds for expressing extreme symptoms compared to participants with secondary education and more (OR = 7.62, 95% CI: 2.57-22.60, p = 0.000), which is a very strong relationship between lesser education and severity in OA symptoms in knees.

Individuals earning between 25,001 to 49,999 had 2.47 times greater odds for extreme symptoms (OR = 2.47, 95% CI: 1.19-5.13, p = 0.015) compared to those earning less than 25,000. This suggests that middle-income participants may face an increased symptom burden. Dependent individuals had 1.78 times greater odds of experiencing severe symptoms (OR = 1.78, 95% CI: 1.01-3.14, p = 0.045) compared to financially independent individuals, highlighting that financial dependence contributes to increased symptoms. Obesity was closely related to both severe and extreme symptoms. Participants with obesity had significantly greater odds for severe (OR = 3.07, 95% CI: 1.61-5.83, p = 0.001) and extreme symptoms (OR = 5.26, 95% CI: 2.65-10.47, p = 0.000). This reaffirms obesity’s well-established relationship with aggravated symptoms for knee OA. Diabetes was also significantly related to symptom severity. Participants with diabetes had 6.05 times greater odds for severe symptoms (OR = 6.05, 95% CI: 2.08-17.53, p = 0.001) and 3.16 times greater odds for extreme symptoms (OR = 3.16, 95% CI: 0.97-10.30, p = 0.055), implying that diabetes aggravates symptoms for knee OA. Participants with CVD had 2.5 times greater odds for extreme symptoms (OR = 2.50, 95% CI: 1.27-4.94, p = 0.000) compared to non-CVD participants, suggesting that cardiovascular illnesses further add to symptom severity in knee OA individuals.

The model classifies 57.4% of the cases correctly, which is a reasonable performance. The AIC and BIC values indicate that the model is reasonably efficient. Multicollinearity was not observed. The results of the regression show that the most persistent and significant predictors of severity are age, obesity, and diabetes. The factors have been proven to remain significant in unadjusted and adjusted models, illustrating that they play a dominant role in predicting severity levels in the WOMAC. Other variables such as gender, education, and CVD also showed significant effects but were more variable across different model adjustments.

## Discussion

The key results of this research indicated substantial correlations between socio-demographic variables, comorbidities, and symptom severity of knee osteoarthritis measured with WOMAC scores.

Age was found to be a determinant of symptom severity in knee OA. Participants over 70 years old had 4.79 fold increased chances of severe symptom experiences compared to 60 to 69 year old participants. This is corroborative of earlier studies that indicated older people reported increased intensity of pain and functional disability related to OA compared to younger individuals [26,27]. Another similar study in Saudi Arabia similarly concluded that age was a better predictor of WOMAC severity than obesity or hypertension. In this research, participants aged ≥50 years had significantly elevated WOMAC index scores, where 15.96% of participants were rated as high-risk with severe knee OA [28].

Gender also played an important role in severity and experience of symptoms of OA of the knee. Female participants of the study were 2.75 times more prone to extreme symptoms compared to their male counterparts, as several studies have concluded that the symptoms of OA of the knee afflicted more women compared to men [29,30]. A study conducted in Saudi Arabia also revealed that 73% of the female patients who had high WOMAC scores also had high clinical OA diagnoses [28].

The educational status contributed significantly to the severity of the symptoms of OA knees. Participants involved in this study who were illiterate had 7.62 times the opportunities of developing extreme symptoms than those who had an educational status of secondary and above. This was evident using a longitudinal study among 1,499 older adults where those without formal qualification had 4.33 more opportunities for severe functional disability [31]. Poorer educational status was also commonly associated with manual laboring occupations [32], which may amplify knee joint loading, with a corresponding increased severity of symptoms.

Income was also a contributing determinant to the severity of symptoms of OA of the knees. Participants who were of low income had 2.47-fold higher odds of severe symptoms. The outcome showed similarity to that of a Korean study, where the lowest-income category experienced higher prevalence of knees OA seen on radiographs (34.4%), compared to the high-income category (11.3%) [33]. Economic factors and severity of symptoms showed that considerations were affecting both function and physical impact of OA of the knees.

Symptom severity was also related to financial independence. Those that were dependent had 1.78 times increased risk of severe knee OA symptom presence than non-dependent participants. This was in accordance with the English Longitudinal Study of Ageing, where financial dependency partly influenced knee OA symptom presence through obesity, with more obesity found in lower-income individuals. A more intense load on the joints and increased pain were the effects of obesity [31].

Participants with obesity were found to have 5.26 times the odds of extreme knee osteoarthritis symptom development. Several studies have illustrated a direct linear association between body mass index (BMI) and increased WOMAC severity. In a cohort of 79 older adult patients with knee OA, class III obesity was linked to an 178% increased WOMAC symptom score compared to individuals with a normal BMI [34]. Additionally, a cohort study in the UK of 1,499 older patients with symptomatic knee OA also reported that 47.4% of patients were obese [31]. Moreover, a study of Chinese women with a BMI of ≥24 kg/m² and the use of oral contraceptive medications likewise reported twice the risk of knee pain [35].

This research also determined that diabetic participants had 6.05 fold more severe knee OA symptoms. Similar research demonstrated that the incidence of type 2 diabetes in older patients having a total knee arthroplasty due to knee OA increased to 16.1% [36]. A population study in Taiwan demonstrated that patients with type 2 diabetes (T2DM) had a 2.75-fold increased risk of developing knee OA compared to non-diabetic patients [37]. This highlights the worsening effect of diabetes in intensifying OA symptoms in the knee.

In addition, participants with CVD suffered 2.5 more extreme knee OA symptoms than those without CVD. Another study reported that 61.5% of the older population with knee OA had hypertension and 43.3% presented with dyslipidemia, two key cardiovascular risk factors [38]. A South Korean study that included 9,514 adults older than 50 years old found that hypertensive patients showed a 26% higher incidence of knee osteoarthritis [39].

## Conclusion

The study provides compelling evidence of the impact of socio-demographic variables and comorbid conditions on the severity of knee osteoarthritis symptomology using WOMAC scores. Increased older age, female sex, low educational attainment, and the presence of obesity, diabetes, and cardiovascular disease were all found to exacerbate the symptomology of osteoarthritis of the knee. The study serves to further support the multifactorial etiology of the disease and its correlation with the social determinants of health. This association serves to underscore the need for a more holistic approach to the treatment of osteoarthritis of the knee that not only deals with the disease’s physical element, but also its socio-economic context.

## Recommendations

It is essential to embrace an integrated healthcare strategy that integrates medical and socio-economic interventions. Programs need to be developed by policymakers to counter the socio-economic determinants of disease that affect an individual’s condition by targeting education, income disparities, and improving healthcare access, particularly for vulnerable groups such as the elderly, women, and poorer and less educated individuals. As these comorbid conditions of obesity, diabetes, and cardiovascular disease substantially affect the severity of osteoarthritis of the knees, there needs to be specific screening for these conditions for particular risk groups.

Early diagnosis and treatment can also slow down the progression of symptoms and provide improved prognosis for the future. There is a requirement to have intense outreach within the community to make people aware of the risk factors related to knee osteoarthritis, especially within disadvantaged areas. Health education programs specifically designed to benefit illiterate people and the poorer population could empower them to take proactive measures to ensure their health and obtain appropriate care in time. Further research will be essential to identify the long-term influence of socio-demographic variables and comorbid conditions upon the progression of knee osteoarthritis. Longitudinal studies have the potential to identify association and influence the development of more efficacious, tailored approaches to managing knee osteoarthritis. The healthcare systems should incorporate integrated care strategies that incorporate physical rehabilitation, obesity and weight loss, diabetes control, and cardiovascular disease monitoring under a unified framework to manage knee osteoarthritis. A multidisciplinary strategy would provide holistic care to patients, enhancing both their physical function and quality of life.

## Supporting information

Informed Consent Form

Questionnaire

## Data Availability

All data produced in the present study are available upon reasonable request to the authors

## Acknowledgment

The authors are grateful to the Dhaka Medical College Hospital (DMCH), National Institute of Traumatology and Orthopaedic Rehabilitation (NITOR), and Bangladesh Institute of Research and Rehabilitation in Diabetes, Endocrine and Metabolic Disorders (BIRDEM) staff and participants for their immense support and coordination throughout the study.

## Supporting information

S1 Appendix. Informed consent form. The written consent form was administered to the participants.

S2 Appendix. Questionnaire. The included questions comprised the questionnaire administered to the participants.

S3 Appendix. Data. This file contains research data.

## Funding Statement

The study did not receive any funding from any agency in the public, commercial, or not-for-profit sectors

## Author Contribution

Conceptualization: Dr. Mohoshina Karim, Md. Delwar Hossen, Tasrima Trisha Ratna

Data curation: Md. Delwar Hossen, Joynal Abedin Imran, Rahnuma Hossain Twasin

Formal analysis: Md. Delwar Hossen, Fatema Priyanka, Shahriar Hasan

Investigation: Md. Delwar Hossen, Joynal Abedin Imran, Rahnuma Hossain Twasin

Methodology: Dr. Mohoshina Karim, Tasrima Trisha Ratna, Md. Delwar Hossen, Fatema Priyanka

Project administration: Dr. Mohoshina Karim, Fatema Priyanka, Umme Salma

Resources: Dr. Ilat-E-Mees Subah, Marzana Afrooj Ria

Software: Fatema Priyanka, Shahriar Hasan

Supervision: Dr. Mohoshina Karim, Dr. Ilat-E-Mees Subah

Validation: Dr. Ilat-E-Mees Subah, Shahriar Hasan, Umme Salma

Visualization: Fatema Priyanka, Joynal Abedin Imran

Writing – original draft: Tasrima Trisha Ratna, Fatema Priyanka, Rahnuma Hossain Twasin

Writing – review & editing: Dr. Mohoshina Karim, Md. Delwar Hossen, Joynal Abedin Imran

## Declaration of Conflict of interest

The authors declare that they have no known competing financial interests or personal relationships that could have appeared to influence the work reported in this paper.

## Notes

### Competing Interest Statement

The authors have declared no competing interest.

### Funding Statement

This study did not receive any funding

