## Supplementary material for "Mapping the intersection of social status and comorbidity in knee osteoarthritis: a WOMAC-based study": Informed Consent Form

Principal Investigator: Dr. Mohoshina Karim

My name is (mention the interviewer’s name).

**Project Goal**:

Congratulations! You have been chosen to participate in this research study. This study is designed to learn about impact of knee osteoarthritis among older individuals, such as yourself.

**Procedures**: I'd like to have an interview with you if you're willing to participate in this study. This will not take more than 15 minutes. We're not going to draw blood, urine, or any other samples from you for this study. I'll tell you some other things about your privacy, rights, and risks or benefits if you're willing to participate in this study. Don't worry, I promise that even if you don't participate in this study, you won't lose any benefits that you usually receive from this hospital.

**Confidentiality:** All the information given by you in this study will be kept confidential and will be used only for academic and research purposes.

**Participant's rights, risks, and benefits:** Grasp that your involvement in this study is purely voluntary and that you may withdraw from the study at will after being enrolled. After being enrolled in this study, you will answer some questions. However, you have the right to refuse to answer any questions in the course of an interview. Further understand that while there is no direct participant in being in this study, results from this study will contribute in the future towards an improved understanding of an older person's health needs for knee osteoarthritis.

**For any further questions, you can reach one of us by calling the above phone numbers.**

**Participant's consent:** I have been informed well about this study. I give my consent consciously and voluntarily to take part in this study.

______________________	__________________

**Signature of the Participant	Date**

**Mobile No.: ___________________**

**_______________________	___________________**

**Signature of researcher**	**Date**
