## Supplementary material for "Mapping the intersection of social status and comorbidity in knee osteoarthritis: a WOMAC-based study": Questionnaire

Serial No……………………………………. Date………………………………….

Name……………………………...

| **Variables** | **Categories** | **Code** |
| --- | --- | --- |
| **Socio-demographic characteristics:** | | |
| a) Age | a) 60 to 69 years b) 70 and above years | 1  2 |
| b) Gender | a) Male b) Female | 1  2 |
| c) Marital status | a) Married b) Unmarried | 1  2 |
| d) Education | a) Illiterate b) Primary c) Secondary and above | 1  2  3 |
| e) Monthly family income | a) 25,000 and below b) 25,001 to 49,999 c) 50,000 and above | 1  2  3 |
| d) Financial independence | a) Independent b) Dependent | 1  2 |
| A. **Disease related factors:** | | |
| 7. Diabetes | a) No b) Yes | 1  2 |
| 8. Obesity | a) No b) Yes | 1  2 |
| 9. Cardiovascular disease | a) No b) Yes | 1  2 |

**Scoring of Osteoarthritis (WOMAC Scale)**

| **Rate your pain when** | **None** | **Slight** | **Moderate** | **Severe** | **Extreme** | **Investigator**  **use only** |
| --- | --- | --- | --- | --- | --- | --- |
| Walking | 0 | 1 | 2 | 3 | 4 |  |
| Climbing stairs | 0 | 1 | 2 | 3 | 4 |  |
| Sleeping at night | 0 | 1 | 2 | 3 | 4 |  |
| Resting | 0 | 1 | 2 | 3 | 4 |  |
| Standing | 0 | 1 | 2 | 3 | 4 |  |
| **Rate your stiffness in the…..** |  |  |  |  |  |  |
| Morning | 0 | 1 | 2 | 3 | 4 |  |
| Evening | 0 | 1 | 2 | 3 | 4 |  |
| **Rate your difficulty when….** |  |  |  |  |  |  |
| Descending Stairs | 0 | 1 | 2 | 3 | 4 |  |
| Ascending stairs | 0 | 1 | 2 | 3 | 4 |  |
| Rising from sitting | 0 | 1 | 2 | 3 | 4 |  |
| Standing | 0 | 1 | 2 | 3 | 4 |  |
| Bending to floor | 0 | 1 | 2 | 3 | 4 |  |
| Walking on even floor | 0 | 1 | 2 | 3 | 4 |  |
| Getting in/out of car | 0 | 1 | 2 | 3 | 4 |  |
| Going shopping | 0 | 1 | 2 | 3 | 4 |  |
| Putting on socks | 0 | 1 | 2 | 3 | 4 |  |
| Rising from bed | 0 | 1 | 2 | 3 | 4 |  |
| Taking off socks | 0 | 1 | 2 | 3 | 4 |  |
| Lying in bed | 0 | 1 | 2 | 3 | 4 |  |
| Getting in/out bath | 0 | 1 | 2 | 3 | 4 |  |
| Sitting | 0 | 1 | 2 | 3 | 4 |  |
| Getting in/out toilet | 0 | 1 | 2 | 3 | 4 |  |
| Doing light domestic duties (cooking, dusting) | 0 | 1 | 2 | 3 | 4 |  |
| Doing heavy domestic duties (moving furniture) | 0 | 1 | 2 | 3 | 4 |  |
| **Total score (out of 96)** | | | | | |  |
